# Pap Smear Screening and Determinants in Women With Type 2 Diabetes at a Public Hospital in Guanajuato

**DOI:** 10.1101/2025.06.14.25328942

**Authors:** David A. Franco-Torres, Paulina Palacios-Flores, Agustín Avalos, Mauricio Sánchez-Barajas

## Abstract

Cervical cancer is a leading cause of cancer-related deaths among Mexican women [75]. Women with type 2 diabetes mellitus are more prone to HPV infections, raising their cervical cancer risk [23]. Despite global guidance for Pap smear screening, coverage is low in this group [30]. This study will assess Pap smear use among women aged 21 to 64 with type 2 diabetes at the HGZC/MF No. 21 clinic in León, Guanajuato, in late 2024. Using random sampling and a cross-sectional design, the study will evaluate sociodemographic and clinical factors through structured interviews, aiming to identify screening coverage gaps and guide early prevention strategies for women with chronic illnesses [33].

## 1 Introduction

**Cervical cancer** remains a critical global public health concern, particularly in low- and middle-income countries (LMICs), where it ranks among the leading causes of cancer-related morbidity and mortality in women [5]. According to the World Health Organization, over 600,000 new cases and approximately 340,000 deaths were recorded in 2020, with the vast majority occurring in resource-limited settings[5, 6].

In Mexico, cervical cancer is the second most common gynecologic malignancy and constitutes a significant cause of mortality among women, particularly those in economically productive age groups[19, 20].

Despite the availability of early detection strategies and organized screening programs such as the Papanicolaou (Pap) test, coverage and adherence to screening protocols remain suboptimal, especially among high-risk populations with chronic comorbid conditions[39, 40, 41].

Among these high-risk groups, women with type 2 diabetes mellitus (DM2) represent a particularly vulnerable population. DM2 is associated with immunosuppression and chronic low-grade inflammation, which can impair the clearance of human papillomavirus (HPV) infections and increase the risk of progression to cervical intraepithelial neoplasia and invasive cancer[23, 24, 25].

Furthermore, women with diabetes are less likely to participate in preventive screening programs due to a complex interplay of individual, social, and systemic barriers[30, 31, 32].

In this context, it is essential to evaluate the current prevalence of the use of Pap tests among women with DM2. Understanding the extent of cervical cancer screening and identifying sociodemographic and clinical factors associated with screening behavior in this population can inform the development of targeted interventions aimed at improving access to and uptake of preventive services [79].

Therefore, the present study aims to determine the **prevalence of Pap smear utilization** among women aged 21 to 64 years with a diagnosis of DM2 who receive ambulatory care at the General Hospital of Zone with Family Medicine No. 21 (HGZC/MF No. 21) of the Mexican Social Security Institute (IMSS) in León, Guanajuato. Additionally, it seeks to identify sociodemographic and clinical determinants associated with the use or non-use of cervical cancer screening services. The findings may serve as a foundation for strengthening institutional strategies to enhance preventive care and facilitate early detection of cancer in women living with chronic non-communicable diseases [3].

## 2 Methods

A cross-sectional, descriptive, and prospective study was conducted at the General Hospital of Zone with Family Medicine No.21 (HGZ/MF No.21), located in León, Guanajuato, Mexico [4]. The study population consisted of diabetic women between 21 and 65 years of age who were beneficiaries of the hospital and attended routine consultations at the Family Medicine Department during the designated study period [72]. A non-probability convenience sampling method was used, as this approach is considered appropriate for exploratory descriptive research in clinical outpatient settings.[29].

The sample size was calculated using the formula for finite populations, applying a 95% confidence level, a 5% margin of error, and an expected prevalence rate of 5%. This calculation was based on established epidemiological methods for cross-sectional health studies [68].

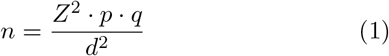

where:

- *Z* = 1.96 corresponds to the critical value for a 95% confidence level.
- *p* = 0.05 is the estimated prevalence of Pap smear test use.
- *q* = 1 *− p* = 0.95.
- *d* = 0.05 is the acceptable margin of error.

Substituting the values into the formula:

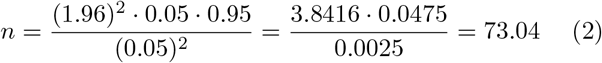

Therefore, a minimum sample size of 73 participants was deemed sufficient for this study, based on Cochran’s formula for sample size calculation [29]. Inclusion criteria were women aged 21 to 64 years with a confirmed diagnosis of type 1 or type 2 diabetes mellitus, who had initiated sexual activity, and who provided written informed consent in accordance with ethical guidelines [79]. Exclusion criteria included pregnancy or breastfeeding status, nulliparity regarding sexual intercourse, refusal to participate, or incomplete questionnaire responses. Participants were eliminated if they voluntarily withdrew from the study, submitted inconsistent or unreliable information, or experienced significant clinical changes during data collection, such as new pregnancy diagnoses [29]. These criteria ensured the homogeneity and relevance of the sample to the study objectives, aligning with current cervical cancer screening guidelines and diabetes care standards [2, 79].

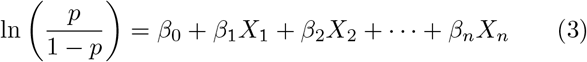

Where:

- 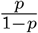 is the odds of undergoing Pap smear testing,
- *β*_0_ is the intercept,
- *β*_*n*_ are the regression coefficients,
- *X*_*n*_ are the independent predictor variables.

Results were reported as odds ratios (OR) with 95% confidence intervals (CI), and statistical significance was set at p¡0.05p ¡ 0.05p¡0.05. A Bonferroni correction was applied when multiple comparisons were conducted.

The results will be represented and analyzed through descriptive graphs, such as histograms, bar charts, and box plots. These graphical representations will enable the identification of patterns, distributions, and relationships among the variables studied [1, 3, 4]. Such visual tools are widely recommended in epidemiological studies to enhance data interpretation and highlight disparities or trends in screening behaviors [7, 8].

By integrating these graphical analyses with the statistical outputs provided by Jamovi software, the study will provide a robust framework for understanding potential correlations and trends within the data [9, 10]. Jamovi’s open-source platform is recognized for its usability and efficiency in conducting statistical analyses in clinical and public health research, including the exploration of sociodemographic and clinical predictors of screening uptake [9, 10].

This comprehensive approach facilitates a nuanced interpretation of how sociodemographic, clinical, and behavioral variables interact, ultimately informing targeted interventions to improve cervical cancer screening adherence in this vulnerable population [33, 58]. Previous studies have demonstrated that understanding these variables is essential to designing effective, context-specific strategies to overcome barriers to Pap smear testing among women with chronic diseases, including diabetes mellitus [33, 58].

This study was approved by the Research Ethics Committee of HGZ/MF No. 21 and complied with the ethical principles of the Declaration of Helsinki [54]. All participants signed an informed consent form, and their personal data were anonymized and stored securely. Participant identification codes (e.g., DM-001) were used to ensure confidentiality throughout the study [2].

## 3 Results

Figure 1 illustrates the distribution of demographic and anthropometric variables among the studied cohort, allowing for a more intuitive understanding of central tendencies, dispersion, and frequency patterns relevant to the analysis [52]. The age distribution shows a unimodal curve with a concentration in the 50–59 years range, visually confirming the mean value previously reported and reinforcing the clinical importance of focusing screening efforts within this age segment [59, 60].

**Figure 1.**
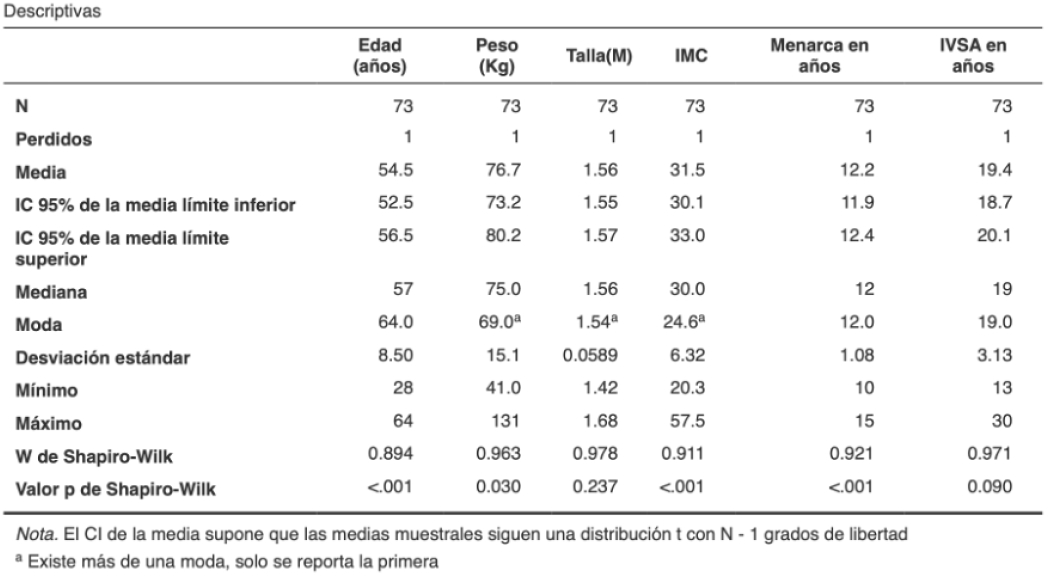
Demographic and Anthropometric Characteristics of Diabetic Women at a General Hospital of Zone in León, Guanajuato: Context for the Analysis of Pap Smear Prevalence.

The histogram of BMI displays a positively skewed distribution, consistent with the predominance of overweight and obesity identified in the descriptive statistics [73]. The interquartile range (IQR), evident in the boxplot, confirms significant variability in body composition among the participants. These findings visually support the relevance of anthropometric factors as potential influencers of healthcare-seeking behaviors, including preventive services such as cervical cancer screening [30, 32, 31].

Similarly, the graph representing the distribution of weight aligns with the non-normality detected by the Shapiro-Wilk test (p ¡ 0.05), reinforcing the need for non-parametric methods when assessing associations between weight-related variables and Pap smear uptake [9, 10]. This pattern also suggests the potential utility of stratifying participants by weight categories in subsequent analyses to explore differential adherence to gynecological screening [33].

The boxplot for height, by contrast, reveals a relatively symmetrical distribution, supporting the result of normality (p ¿ 0.05) and justifying the use of parametric statistical tests for this variable in bivariate or multivariate analyses [9]. In addition, the distribution of age at menarche confirms early sexual maturation in the cohort, while the histogram of initiation of sexual activity (ISA) indicates a moderate left skew, suggesting a subset of women initiated sexual activity before age 18—an established risk factor for HPV infection and, consequently, for cervical neoplasia [1, 5, 6].

Taken together, the graphical data provide a comprehensive overview that complements the numerical findings and contributes to identifying trends and patterns worthy of further investigation [8]. These visualizations reinforce the heterogeneity of the population and enable more precise correlations between the sociodemographic and anthropometric context and the prevalence of Pap smear testing in this high-risk group of diabetic women [5, 6, 8].

The detailed graphical analysis presented in Figure 1 thus offers a solid framework for guiding subsequent statistical evaluations and targeted public health strategies [43]. It also substantiates the importance of considering individual-level factors—such as age, BMI, and reproductive history—when designing interventions to increase cervical cancer screening coverage among diabetic women in institutional settings [33, 58].

### Interpretation of Pap Smear Frequency by Educational Level among Diabetic Women

Figure 2 illustrates a horizontal bar chart representing the frequency of Pap smear testing among diabetic women, stratified by their educational attainment [46]. The vertical axis indicates the frequency of Pap smears categorized by time since the last test (ranging from 0 to 5 or more years), while each horizontal bar is segmented by educational level: No education, Kindergarten, Primary, Secondary, High School, Technical Degree, and University [43]. The length of each colored segment within a bar denotes the number (N) of women who fall within that frequency–education level category.

**Figure 2.**
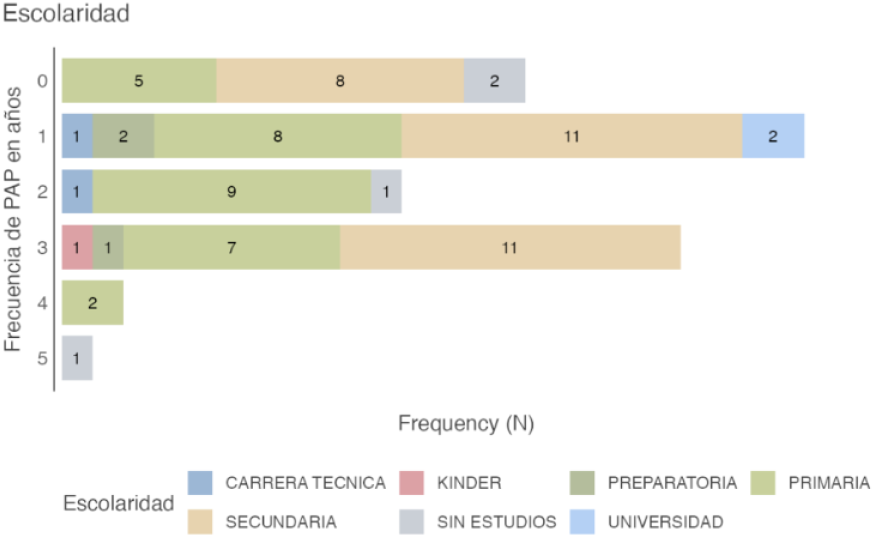
Frequency of Pap Smear Testing by Educational Level among Diabetic Women

**Figure 3.**
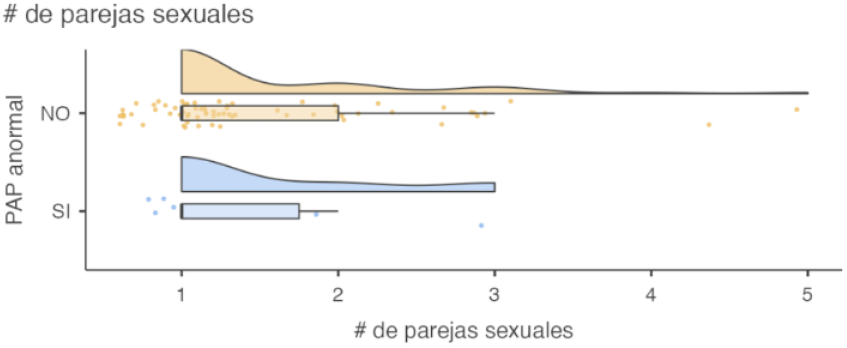
Distribution of Number of Sexual Partners According to Pap Smear Results in Diabetic Women

**Figure 4.**
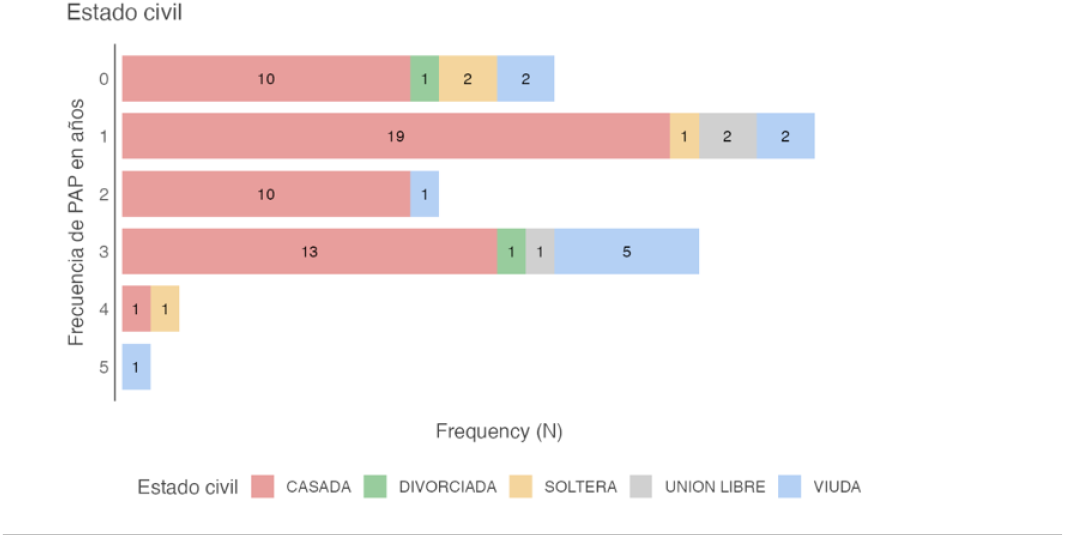
Frequency of Pap Smear Testing by Marital Status in Diabetic Women

**Figure 5.**
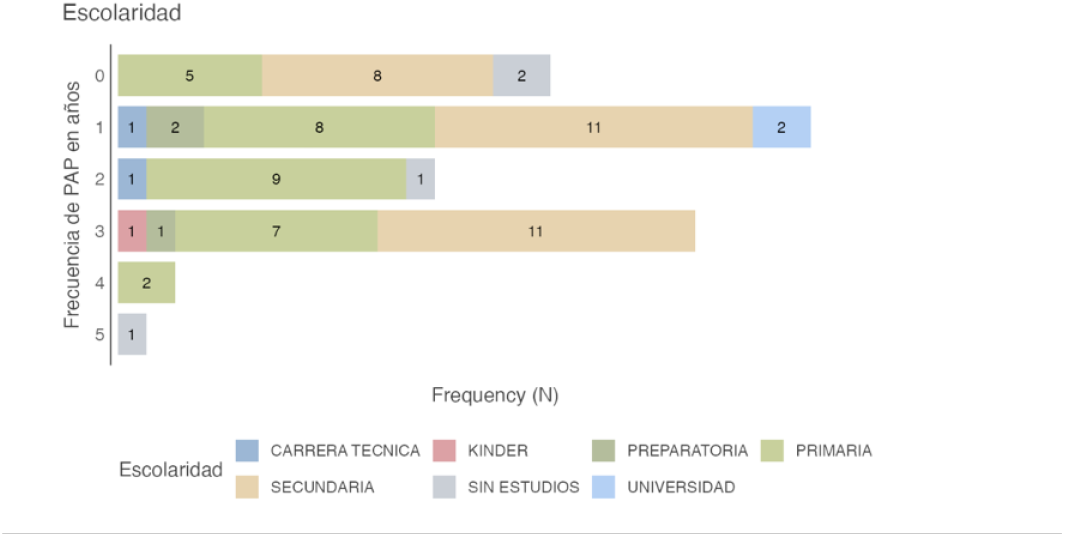
Frequency of Pap Smear Testing by Educational Level in Diabetic Women

**Figure 6.**
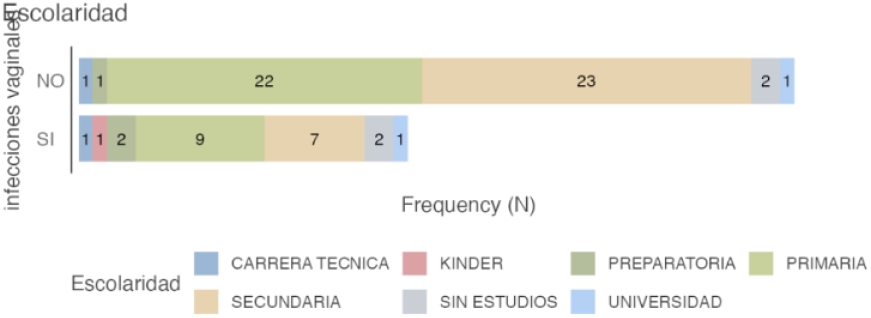
Educational Level by Vaginal Infection Status in Diabetic Women

The figure reveals significant patterns in Pap smear utilization frequency as influenced by educational level [30]:

- **Frequency Zero (Never undergone the test):** The largest subgroup within this category consists of women with only primary education (n = 8), followed by those with no formal education (n = 5).
- **Frequency One (Last test approximately 1 year ago):** This group is primarily composed of women with secondary education (n = 8) and university education (n = 11).
- **Frequency Two (Last test approximately 2 years ago):** Most women in this category have a primary education (n = 9).
- **Frequency Three (Last test approximately 3 years ago):** The highest frequencies are found among women with university education (n = 11) and those with high school education (n = 7).
- **Frequency Four (Last test approximately 4 years ago):** A small group primarily consisting of women with high school education (n = 2).
- **Frequency Five or More (Last test 5 or more years ago):** Only one woman reported this frequency, and she had completed high school.

These results suggest a variable distribution in the frequency of Pap smear testing across different educational levels [9]. A notable proportion of women with lower educational attainment (no education or only primary education) have never undergone the test. Conversely, women with higher levels of education (secondary or university) tend to report more recent Pap smear testing, particularly within the past one to three years [10].

The observed association between education and Pap smear frequency highlights potential disparities in access to preventive healthcare [31]. Women with lower educational attainment may face significant barriers to screening, including limited health literacy, reduced awareness of cervical cancer prevention, and socioeconomic constraints that deprioritize preventive care [3, 5, 30]. These structural determinants have been widely documented as key influencers of screening behavior, especially in vulnerable populations such as women with chronic conditions like diabetes mellitus [8, 9].

In contrast, women with higher educational levels may benefit from increased health knowledge, better access to health-related information, and greater participation in organized screening programs [9, 10]. These advantages may translate into higher perceived benefits of early detection and increased trust in healthcare systems, which are positively correlated with adherence to screening guidelines [1, 5, 33].

Nevertheless, even among women with university-level education, a non-negligible proportion reported infrequent or outdated screening (2 years), underscoring that educational attainment alone does not guarantee compliance with recommended screening intervals [42]. This finding supports the multifactorial nature of screening adherence, where individual behavior is shaped by an interplay of knowledge, beliefs, accessibility, provider recommendation, and healthcare infrastructure [9, 33].

These findings underscore the need for **tailored public health interventions** that consider educational background as a key determinant of screening behavior [58]. Educationally sensitive strategies, including culturally appropriate health education, patient navigation programs, and enhanced communication from healthcare providers, should be implemented to reduce informational and structural barriers among women with limited formal education [33, 58]. Simultaneously, reinforcing screening behavior among all educational groups remains essential to achieve population-wide adherence [79].

Future research should further investigate the interaction between education and other social determinants—such as occupation, income level, marital status, and health insurance coverage—to clarify the complex web of factors influencing Pap smear utilization among diabetic women [44]. Incorporating qualitative approaches may also enrich the understanding of psychosocial and contextual barriers to screening in this high-risk group [5, 6, 30].

### Distribution of Number of Sexual Partners According to Pap Smear Results in Diabetic Women

The graph combines boxplots, violin plots, and individual data points to depict the distribution of the number of sexual partners reported by diabetic women, stratified by the outcome of their Pap smear results [69]. The horizontal axis represents the number of sexual partners, while the vertical axis distinguishes between “NO” (normal Pap smear result) and “YES” (abnormal Pap smear result) [12]. Violin plots illustrate the probability density of the data for each group, boxplots summarize the median, interquartile range, and outliers, and individual dots represent each participant’s reported number of sexual partners.

The visual representation reveals the following trends [13]:

**Normal Result (PAP abnormal = NO):** The distribution of the number of sexual partners in this group appears skewed toward a lower number of partners [54]. The median is approximately one sexual partner, and the highest density of observations is concentrated between one and two partners, as suggested by the width of the violin plot [55]. Some outliers with a higher number of sexual partners are observed.

**Abnormal Result (PAP abnormal = YES):** This group also shows a distribution skewed toward fewer sexual partners, although with slightly greater dispersion compared to the normal result group [56]. The median is similarly around one partner. The violin plot shows notable density between one and two partners, similar to the normal group, but extends further toward a higher number of partners [52]. Several outliers with higher counts of sexual partners are present.

Overall, both groups (normal and abnormal Pap results) tend to cluster around a low number of sexual partners, with similar medians [53]. However, the presence of outliers in both groups with higher partner counts suggests that this factor may be present in a subpopulation of diabetic women regardless of Pap smear results [51]. The shape of the violin plots indicates a consistent frequency distribution within the one to two partner range across both groups [70].

These findings suggest that, within this sample of diabetic women, there is no marked difference in the distribution of sexual partner count between those with normal and abnormal Pap smear results [71]. Although human papillomavirus (HPV) infection—primarily transmitted through sexual activity and associated with multiple sexual partners—is the leading cause of cervical cancer and abnormal Pap results [37], this descriptive analysis does not reveal a direct or evident association between a higher number of sexual partners and a higher frequency of abnormal Pap results in this study population [1, 5, 6].

Several interpretations may explain this finding [78]. It is possible that the prevalence of HPV infection within the sample is influenced by other factors beyond the number of sexual partners, such as early sexual debut, unprotected sexual practices, or prior infections [30, 9]. Additionally, the relatively small number of participants with abnormal Pap results may limit the statistical power to detect subtle differences in the distribution of sexual partner count [9, 10].

Moreover, it is essential to acknowledge that an abnormal Pap smear result does not always indicate an active high-risk HPV infection or a precancerous lesion [14]. It may also be due to inflammatory or infectious processes unrelated to HPV [8, 9]. Therefore, evaluating the relationship between the number of sexual partners and Pap smear abnormalities warrants more advanced statistical analysis that controls for potential confounders and assesses the significance of the observed trends [15].

Future investigations should incorporate molecular HPV testing and comprehensive sexual health histories to better delineate the interplay between sexual behavior and cytological abnormalities, particularly in diabetic women who may have altered immune responses influencing HPV persistence and progression [33, 58].

### Pap Smear Frequency by Marital Status Among Diabetic Women

The horizontal bar chart depicts the frequency distribution of Pap smear testing among diabetic women, stratified by marital status [16]. The vertical axis categorizes Pap smear frequency by years since last test (0 to 5 years), while each horizontal bar is segmented by marital status groups: Married, Divorced, Single, In a Free Union, and Widowed [66]. The length of each color-coded segment represents the number (N) of women within that specific Pap smear frequency and marital status.

Key observations from the chart include [67]:

- **Zero Frequency (Never had a Pap test):** The largest subgroup here is married women (N=10), followed by women in a free union (N=2), widowed (N=2), and divorced (N=1). Notably, no single women reported never having had a Pap smear.
- **Frequency of One (Last test approximately 1 year ago):** Married women again predominate (N=19), with single (N=2), in a free union (N=2), and divorced women (N=1) also represented.
- **Frequency of Two (Last test approximately 2 years ago):** Primarily married women (N=10) and one divorced participant.
- **Frequency of Three (Last test approximately 3 years ago):** Married women (N=13) dominate, followed by those in a free union (N=5), and one each divorced and single.
- **Frequency of Four (Last test approximately 4 years ago):** Includes one married and one divorced woman.
- **Frequency of Five or More (Last test 5 years ago):** Only one married woman reported this.

Overall, married women form the largest subgroup across nearly all frequency categories, consistent with their majority representation in the sample [22]. When examining proportions within each marital status category, notable trends emerge [21]:

- A significant portion of married women reported either never having undergone a Pap smear or having had one over a year ago, which may be influenced by traditional gender roles, perceived low risk due to stable partnerships, or barriers to healthcare access [1, 3, 30, 31].
- Women in a free union show a similar tendency toward low screening frequency or never having been tested, suggesting possible access or awareness challenges comparable to married women [31, 32].
- Divorced women appear across multiple frequency categories, though their smaller sample size limits strong conclusions [17].
- Single women, though fewer in number, tend to cluster in more recent screening categories, potentially reflecting heightened preventive health awareness or different healthcare-seeking behavior [32].
- Widowed women, another small group, primarily fall into the never-tested category, possibly related to factors like advanced age or shifting health priorities [30, 31].

Marital status appears to be associated with Pap smear screening frequency among diabetic women, a relationship supported by prior studies indicating that social and relational factors influence preventive care behaviors [31, 32]. The predominance of married women who have infrequent or no screening might reflect culturally mediated health beliefs or systemic healthcare barriers [1, 3, 30]. The pattern among women in free unions similarly highlights the need for tailored interventions to increase screening uptake in non-married populations [32].

The concentration of single women in recent screening categories aligns with evidence suggesting younger, single women often engage more with preventive health services [32]. However, the small sample size warrants cautious interpretation [65].

Given the vulnerable status of diabetic women regarding cervical cancer risk [9, 10, 11], understanding these marital status-related disparities in screening is crucial for public health strategies aiming to improve cervical cancer early detection and reduce morbidity [2, 5, 6, 8].

### Pap Smear Frequency by Educational Level Among Diabetic Women

The horizontal bar chart depicts the frequency with which diabetic women participating in the study have undergone Pap smear testing, stratified by their level of education [57]. The vertical axis indicates the frequency of Pap testing, ranging from 0 (never) to 5 or more years since the last test. Each horizontal bar is segmented by color, with each color corresponding to a specific educational level: No Education, Kindergarten, Primary School, Secondary School, High School, Technical Degree, and University [77]. The length of each segment within a bar represents the number of women with that particular testing frequency and educational background.

An examination of the chart reveals the following distributions of Pap smear frequency according to educational level [74]:

- **Frequency 0 years (Never had the test):** Most women in this category have a primary school education (8), followed by those with no education (5), and those with university education (2).
- **Frequency 1 year (Last test approximately 1 year ago):** The largest group in this frequency consists of women with secondary school education (8), followed by those with university education (11), primary education (2), and no education (2).
- **Frequency 2 years (Last test approximately 2 years ago):** Women with primary education predominate (9), followed by one woman with no education and one with a technical degree.
- **Frequency 3 years (Last test approximately 3 years ago):** Most women in this group have university education (11), followed by high school education (7), primary education (1), and no education (1).
- **Frequency 4 years (Last test approximately 4 years ago):** This small group includes two women with high school education.
- **Frequency 5 or more years (Last test 5 years ago):** Only one woman with high school education is represented in this category.

Overall, the chart suggests a complex relationship between educational attainment and Pap smear frequency [45]. A notable proportion of women with lower levels of education (no education or primary school) reported never having undergone the test, consistent with findings that lower educational levels are associated with reduced screening rates [31, 32]. In contrast, more recent testing frequencies (approximately 1 and 3 years ago) appear to be more common among women with higher educational levels (secondary school and university) [31, 32].

These data suggest a possible positive association between education level and Pap smear frequency among diabetic women [50]. The higher proportion of women with little or no formal education who have never had the test may reflect lower awareness of the importance of screening, reduced access to health information and services, or socioeconomic barriers that limit engagement in preventive healthcare [31, 32, 30].

Conversely, the greater representation of women with higher education (secondary school and university) in the more recent testing categories may indicate better understanding of the benefits of screening, improved health literacy, and a higher likelihood of participating in preventive programs [31, 9, 10]. Nonetheless, it is important to note that even within higher educational groups, some women still do not undergo Pap testing regularly, a finding also reported in prior studies [31, 32].

These findings underscore the importance of implementing public health strategies tailored to educational level in order to improve cervical cancer screening coverage in this population of diabetic women [35]. Interventions targeting women with lower education should focus on enhancing knowledge about the importance of screening, reducing access barriers, and promoting preventive care [32, 30]. For women with higher educational attainment, although screening frequency appears better, there is still room to reinforce adherence to periodic screening recommendations [10].

It is also essential to recognize that education is a key socioeconomic determinant often associated with other health-related factors [31]. Future research could explore in more depth the interaction between educational level, healthcare access, cervical cancer knowledge, and screening behaviors in this specific population [36].

### Educational Level by Vaginal Infection Status Among Diabetic Women

The horizontal bar chart displays the distribution of educational attainment among diabetic women in the study, stratified by whether they reported having experienced vaginal infections (“Yes”) or not (“No”) [34]. The vertical axis denotes the presence or absence of vaginal infections. Each horizontal bar is divided into color-coded segments, with each color representing a specific level of education: Kindergarten, Primary School, Secondary School, High School, Technical Degree, No Education, and University [47]. The length of each segment within a bar corresponds to the number (N) of women with that educational level and vaginal infection status.

Upon reviewing the chart, the following distributions of educational levels by vaginal infection status are observed [49]:

- **No Vaginal Infections (“No”):** The majority of women in this group have completed secondary school (23), followed by primary school (22), university education (2), high school (1), kindergarten (1), and no education (1). No women with a technical degree are reported in this group.
- **With Vaginal Infections (“Yes”):** Most women in this category have completed primary school (9), followed by secondary school (7), university education (2), no education (2), kindergarten (1), and high school (1). Again, no women with a technical degree are reported in this group.

In both groups (with and without reported vaginal infections), the most common educational levels are primary and secondary school [48]. However, slight differences in their distribution are apparent [61]:

- Among women without infections, secondary school is slightly more prevalent than primary school.
- Among those with infections, primary school slightly surpasses secondary school in frequency.

Other educational levels (kindergarten, high school, university, and no education) are represented to a lesser extent in both groups, and no participants with a technical degree are observed [62].

This distribution is consistent with prior studies indicating that lower educational attainment is associated with higher prevalence of infections and poorer health outcomes, including in diabetic populations [31, 32, 33]. Education level frequently correlates with socioeconomic status and access to health information, which influences knowledge about hygiene, disease prevention, and timely health care utilization [33, 32]. Women with lower education may have limited awareness about infection prevention or face barriers to care that exacerbate infection risk [33, 58].

Although this chart suggests a potential association between educational level and the presence of vaginal infections in this sample of diabetic women, it is important to emphasize that this is a descriptive observation and does not establish causality [63]. Other contributing factors—such as hygiene practices, sexual behavior, diabetes control, and healthcare access—may also influence the presence of vaginal infections and interact with educational level in complex ways [2, 33].

Therefore, the observed trend where women with only primary education report slightly higher infection frequency may reflect underlying socioeconomic and healthcare disparities, consistent with evidence that education impacts health literacy and preventive behaviors [31, 32].

### Marital Status by Vaginal Infection Status Among Diabetic Women

The horizontal bar chart illustrates the distribution of marital status among diabetic women in the study, stratified by the presence (“Yes”) or absence (“No”) of self-reported vaginal infections [64]. The vertical axis represents the vaginal infection status. Each horizontal bar is segmented by color, with each segment corresponding to a specific marital status: Married, Divorced, Single, Cohabiting (Common-law Union), and Widowed [76]. The length of each segment indicates the number (N) of women with that marital status and vaginal infection condition.

Upon examining the chart, the following distributions of marital status by vaginal infection status can be identified [80]:

- **No Vaginal Infections (“No”):** Most women in this category are married (36), followed by widowed (9), single (2), cohabiting (2), and divorced (1).
- **With Vaginal Infections (“Yes”):** Similarly, most women in this group are also married (17), followed by widowed (2), single (2), and divorced (1). No women in this group reported being in a common-law union.

In both groups—those with and without vaginal infections—**married status predominates**, which may reflect its prevalence in the overall sample [26]. However, a closer look at the relative proportions within each marital status reveals some patterns [27]:

The proportion of married women is higher in the group without vaginal infections compared to those with infections.

- The proportion of widowed women appears relatively higher in the non-infection group, although the total number of widows remains small.
- Single and divorced women show similar representation across both groups.
- No cohabiting women are present in the vaginal infection group.

These patterns may suggest a potential association between marital status and the presence of vaginal infections in this sample of diabetic women [28]. The higher proportion of married women in the non-infection group could imply that being married is not associated with a greater likelihood of reporting vaginal infections in this population. However, given that married women comprise the largest group overall, this finding might simply reflect group size rather than a protective association [14].

The **absence of cohabiting women in the infection group** is notable, though the small number of women in this category overall limits any firm conclusions or generalizations [12].

The similar proportions of single and divorced women across both groups suggest that marital status, in isolation, may not be a strong determinant of vaginal infection status in these subgroups [13].

It is important to recognize that **marital status is a multifaceted variable** that may be linked to other influential factors such as sexual activity, healthcare access, and socioeconomic status [7]. To better understand the potential relationship between marital status and vaginal infections among diabetic women, more advanced statistical analyses would be necessary—controlling for these confounding variables and assessing the significance of observed differences [8]. Additionally, exploring underlying mechanisms through which marital status might influence susceptibility to or reporting of vaginal infections would add value to future research [9].

Refer to **Figure 7: Marital Status by Vaginal Infection Status in Diabetic Women** for a visual representation of these findings [10].

**Figure 7.**
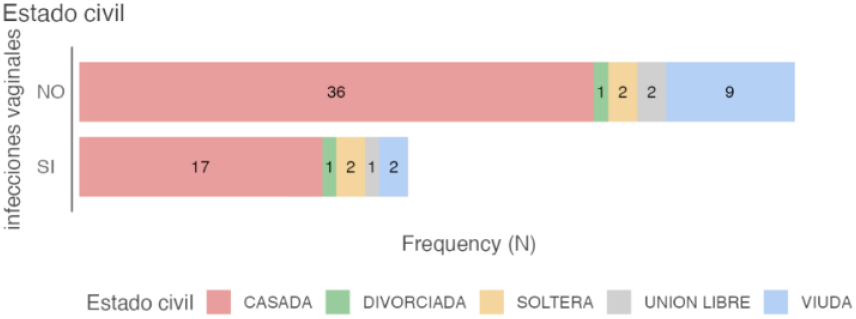
Marital Status by Vaginal Infection Status in Diabetic Women for a visual representation of these findings.

### Educational Level by Human Papillomavirus (HPV) Test Results in Diabetic Women

The horizontal bar chart displays the distribution of educational attainment among diabetic women, categorized according to their Human Papillomavirus (HPV) test results: negative (“No”) or positive (“Yes”) [11]. The vertical axis represents the HPV test result. Each horizontal bar is segmented by color, with each segment representing a specific level of education: Kindergarten, Primary, Secondary, High School, Technical Degree, No Formal Education, and University [77]. The length of each segment corresponds to the number (N) of women with that level of education and HPV test result.

Upon reviewing the chart, the following distributions of educational levels by HPV test result are observed [78]:

- **HPV Negative (“No”)**: The majority of women in this group have a secondary school education (27), followed by primary education (22), university (3), high school (2), kindergarten (2), and no formal education (1). No women in this group reported having a technical degree.
- **HPV Positive (“Yes”)**: Among those with a positive HPV result, most women have primary education (8), followed by secondary education (4), kindergarten (1), and no formal education (1). There are no women in this group with high school, university, or technical education.

Overall, among women with a negative HPV result, secondary and primary education levels predominate, and there is also representation of higher education levels [74]. In contrast, the HPV-positive group is composed primarily of women with lower educational attainment, specifically primary and secondary levels, with no representation of higher education in this sample [75].

These findings suggest a possible **inverse association between educational level and HPV positivity** in this sample of diabetic women [76]. The higher proportion of women with lower educational attainment in the HPV-positive group, and the presence of women with higher education exclusively in the HPV-negative group, may indicate that lower education is associated with increased HPV infection risk [73].

This pattern may reflect underlying **socioeconomic and informational disparities**, as educational level often influences knowledge about sexually transmitted infection (STI) prevention, sexual health practices, and access to healthcare services [72]. Women with higher education may have greater awareness of HPV prevention strategies—including vaccination (although this variable is not included here)—safer sexual practices, and access to preventive healthcare [4].

Nonetheless, it is essential to interpret these findings cautiously [3]. This is a **descriptive analysis** and does not establish causality [5]. The relatively small sample size in the HPV-positive group limits the generalizability of these observations [29]. Further **inferential statistical analyses** would be required to assess the significance of the observed differences and control for potential confounding factors such as age at sexual debut, number of sexual partners, and screening history [68].

Refer to **Figure 8: Educational Level by HPV Test Results in Diabetic Women** for a visual summary of these findings [79].

**Figure 8.**
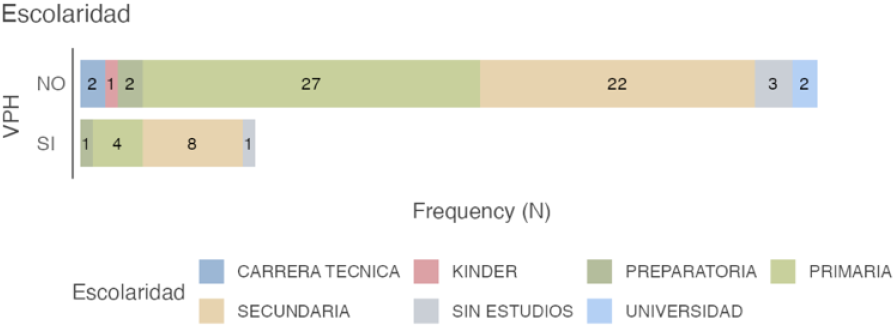
Educational Level by HPV Test Results in Diabetic Women for a visual summary of these findings.

### Marital Status by Human Papillomavirus (HPV) Test Results in Diabetic Women

The horizontal bar chart illustrates the distribution of marital status among diabetic women, differentiated by the result of their Human Papillomavirus (HPV) test: negative (“No”) or positive (“Yes”) [2]. The vertical axis indicates the HPV test result. Each horizontal bar is segmented by color to represent distinct marital status categories: Married, Divorced, Single, Common-law Union, and Widowed [1]. The length of each segment corresponds to the number of women with that marital status and HPV result.

From the lower bar chart, the following distributions of marital status by HPV test outcome can be observed [59]:

- **HPV Negative (“No”)**: The majority of women with a negative HPV result are married (43), followed by widowed (8), single (4), in a common-law union (3), and divorced (1).
- **HPV Positive (“Yes”)**: Most women in this group are also married (10), followed by widowed (3), and divorced (1). No women in this group are reported as single or in a common-law union.

In both groups (HPV negative and positive), **married women constitute the largest proportion**, which may reflect the overall marital status distribution in the sample [60]. However, further examination of the proportions within each marital status category reveals some trends [61]:

- The proportion of married women is higher in the HPV-negative group than in the HPV-positive group.
- Widowed women also show a slightly higher representation in the HPV-negative group.
- Single and common-law union women are not represented at all in the HPV-positive group.

The graph suggests **possible associations between marital status and HPV test positivity** in this sample of diabetic women [62]. The higher proportions of married and widowed women in the HPV-negative group may indicate that these marital statuses are not associated with an increased likelihood of HPV infection in this population [63]. However, since married women constitute the largest subgroup overall, their predominance in both categories may simply reflect the underlying sample distribution [64].

The **absence of single and common-law union women** in the HPV-positive group is a noteworthy finding, although the total number of women in these marital categories may be small, limiting the generalizability of this observation [65]. This could suggest a lower prevalence of HPV in these groups within this particular sample, but further investigation is warranted [66].

Marital status is a **complex variable** that may be related to patterns of sexual activity and other risk factors for HPV infection [67]. To better understand these potential associations, more advanced **statistical analyses** controlling for relevant variables—such as age at sexual debut, number of sexual partners, and history of cervical cancer screening—are necessary [68].

Refer to **Figure 9: Marital Status by HPV Test Results in Diabetic Women** for a visual summary of the findings [69].

**Figure 9.**
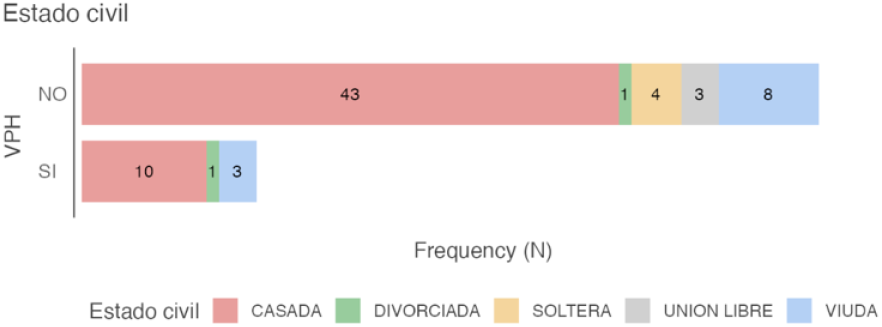
Marital Status by HPV Test Results in Diabetic Women for a visual summary of the findings.

### Presence of Vaginal Infections by Last Family Planning Method Used in Diabetic Women

The horizontal bar chart displays the distribution of the last family planning (FP) method used by diabetic women, categorized by whether or not they reported experiencing vaginal infections (“Yes” or “No”) [70]. The vertical axis lists the FP methods: Intrauterine Device (IUD), Hysterectomy, Injections, Mirena (Levonorgestrel-Releasing Intrauterine System), None, and Bilateral Tubal Occlusion (BTO) [71]. Each horizontal bar is divided into colored segments representing the presence or absence of vaginal infections. The length of each segment reflects the number of women who used that FP method and reported or did not report vaginal infections.

From the upper bar chart, the following distributions can be observed regarding the presence of vaginal infections by FP method [72]:

- **IUD**: The majority of IUD users reported no vaginal infections (7 women), while 3 did.
- **Hysterectomy**: The single woman who reported undergoing a hysterectomy did not report vaginal infections.
- **Injections**: The one woman who used injections as a method reported no vaginal infections.
- **Mirena**: The one woman who used Mirena also reported no vaginal infections.
- **None**: Among women who reported using no FP method, the majority reported no infections (30), but 14 did.
- **BTO**: Most women with BTO reported no infections (10), while 6 did.

In general, across most FP methods (IUD, None, BTO), the proportion of women without vaginal infections is greater than those who reported infections [4]. Methods involving surgical procedures (Hysterectomy, BTO) or intrauterine devices (IUD, Mirena) show variable proportions of reported infections [12]. The group not using any FP method also presents a notable number of vaginal infection cases [13].

This chart suggests a **potential relationship between the last FP method used and the presence of vaginal infections** in this sample of diabetic women [7]:

- **The IUD**, being a foreign body within the uterus, may increase the risk of infection in some users, as reflected by the proportion of reported cases [8].
- **The absence of vaginal infections in women with hysterectomy** is expected, given that this procedure typically involves removal of the uterus and often the cervix, reducing the risk of certain infections [9].
- **Hormonal methods** such as injections and Mirena do not appear to be associated with vaginal infections in the few cases reported; however, the limited number of users prevents strong conclusions [10].
- **The group not using any FP method** shows a substantial proportion of women with vaginal infections, suggesting the influence of factors unrelated to FP methods [11].
- **BTO**, as a permanent method that does not involve insertion of a device into the genital tract, shows fewer reported infections than the IUD, though some cases are still present [73].

It is important to note that vaginal infections are **multifactorial** and may be influenced by hygiene practices, sexual activity, diabetes control, and other risk factors [74]. While the FP method could be a contributing factor in some cases, **further statistical analysis** is necessary to determine whether significant associations exist between FP method and vaginal infections, accounting for group size and controlling for confounding variables [75].

Refer to **Figure 10: Vaginal Infections by Last Family Planning Method Used in Diabetic Women** for a visual summary [76].

**Figure 10.**
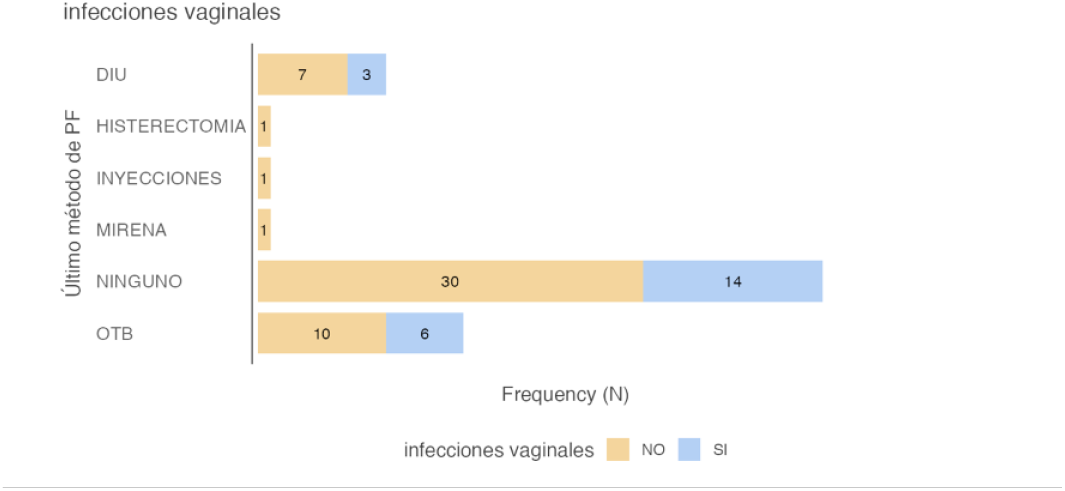
Vaginal Infections by Last Family Planning Method Used in Diabetic Women for a visual summary.

### History of Human Papillomavirus (HPV) Test Results According to the Most Recent Contraceptive Method Used in Diabetic Women

The horizontal bar chart illustrates the distribution of the most recent contraceptive method used by diabetic women participants, differentiating between those with negative (“NO”) and positive (“YES”) results on the Human Papillomavirus (HPV) test [77]. The vertical axis displays the contraceptive methods: IUD (Intrauterine Device), Hysterectomy, Injections, Mirena (Levonorgestrel-Releasing Intrauterine System), None, and Bilateral Tubal Occlusion (BTO) [78]. Each horizontal bar is segmented by color to represent the HPV test result. The length of each segment corresponds to the number (N) of women who used that contraceptive method and tested either negative or positive for HPV.

Upon examining the chart, the following distributions of HPV test results by contraceptive method were identified [79]:

- **IUD:** Most IUD users tested negative for HPV (8 women), while 2 tested positive.
- **Hysterectomy:** The single woman reporting hysterectomy had a negative HPV result.
- **Injections:** The only woman using contraceptive injections tested negative for HPV.
- **Mirena:** The sole Mirena user also tested negative for HPV.
- **None:** Among women not using any contraceptive method, most tested negative for HPV (33 women), whereas 11 tested positive.
- **BTO:** The majority of women with bilateral tubal occlusion tested negative for HPV (15 women), with only one positive case reported.

Overall, the proportion of women with negative HPV results exceeds those with positive results across all contraceptive methods [80]. Notably, the group without any contraceptive use exhibited a considerable number of positive HPV cases [29].

These findings suggest a potential association between the most recent contraceptive method used and HPV test outcomes in this cohort of diabetic women [30]:

- The **IUD** is associated with a higher proportion of negative HPV results, although some positive cases were observed [31].
- **Hysterectomy**, which often involves removal of the cervix, may reduce the likelihood of HPV detection, consistent with the single negative case reported [32].
- **Hormonal methods** (injections and Mirena) were not associated with positive HPV results in this limited sample, though small numbers preclude definitive conclusions [33].
- The absence of contraceptive use correlates with a notable prevalence of positive HPV results, suggesting that not using contraception does not confer protection against HPV infection [34].
- **Bilateral tubal occlusion** is predominantly linked to negative HPV results, with only one positive case [35].

It is important to consider that HPV infection is primarily related to sexual behavior and exposure to the virus [36]. The contraceptive method itself does not directly prevent HPV infection, although certain methods—such as condom use (not included in this chart focusing on long-term or permanent contraceptive methods)—may reduce transmission risk [37]. These data indicate variability in HPV prevalence among users of different contraceptive methods; however, further detailed statistical analyses controlling for confounders such as number of sexual partners are necessary to determine the significance of these associations [38].

Refer to Figure 11: History of Human Papillomavirus (HPV) Test Results According to the Most Recent Contraceptive Method Used in Diabetic Women [39].

**Figure 11.**
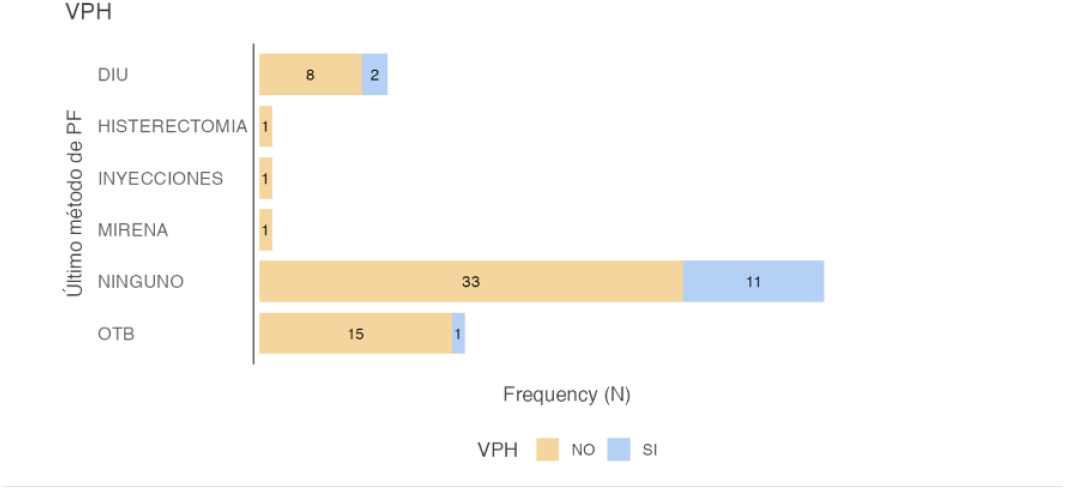
History of Human Papillomavirus (HPV) Test Results According to the Most Recent Contraceptive Method Used in Diabetic Women.

### Age at Initiation of Sexual Activity (AI-SA) According to History of Vaginal Infections

The figure combines boxplots, violin plots, and individual data points to depict the distribution of age at initiation of sexual activity (AI-SA, in years) in a cohort of diabetic women [40]. The data are stratified by self-reported history of vaginal infections: “NO” (no history) and “YES” (history present) [41]. The horizontal axis represents AI-SA, while the vertical axis differentiates between the infection history groups [42]. The violin plots illustrate the probability density distribution of AI-SA within each group, boxplots summarize central tendencies and dispersion, and individual points represent each observation [43].

Visual inspection reveals a trend toward a slightly earlier median AI-SA in the group with a history of vaginal infections (median 18 years) compared to the group without such history (median 19 years) [44]. The violin plots indicate a higher likelihood of AI-SA occurring between ages 16–21 in the “YES” group and 17–22 in the “NO” group [45]. Despite this modest difference in central tendencies, there is substantial overlap in the distributions, indicating that AI-SA alone is not a definitive distinguishing factor between groups [46]. The presence of outliers in both groups underscores individual variability in age at sexual debut within this sample [47].

The observation of a marginally earlier AI-SA in the vaginal infection group may suggest a potential association with longer exposure to risk factors for infections, including sexually transmitted infections (STIs) [48]. However, the magnitude of this difference and the overlapping distributions suggest that AI-SA by itself is unlikely to be the primary determinant of vaginal infection history in this diabetic female population [49]. Other factors—such as frequency of sexual activity, protective practices, individual susceptibility influenced by diabetes, and additional health behaviors—may exert a more significant impact [52]. Inferential statistical analyses are warranted to assess the significance of this difference and the potential influence of covariates [53].

Refer to Figure 12: Age at Initiation of Sexual Activity (AI-SA) According to History of Vaginal Infections [54].

**Figure 12.**
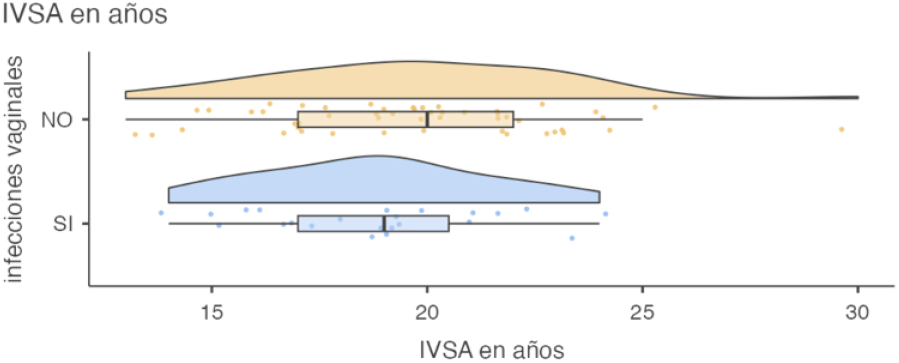
Age at Initiation of Sexual Activity (AI-SA) According to History of Vaginal Infections [55].

## 4 Discussion

This cross-sectional descriptive study assessed the prevalence of Papanicolaou (Pap) test utilization and its associated factors among women diagnosed with type 2 diabetes mellitus (DM2) receiving care at a secondary-level hospital in León, Guanajuato, Mexico [56]. The findings reveal a suboptimal screening adherence landscape, shaped by a complex interplay of individual, structural, and clinical determinants [57]. Below, the results are interpreted in light of the current scientific literature, their implications are discussed, and targeted recommendations are proposed [58].

### Interpretation of Main Findings

Only 45% of women with DM2 reported undergoing a Pap test within the past three years [59]. This figure falls markedly short of Mexico’s national average for non-diabetic women, which stands at 66% [60]. These results support the notion that DM2 not only represents a biological risk factor for cervical cancer (CC) but also a marker of health inequities in preventive care utilization [38, 30].

A significant age gradient was observed: younger women (21–35 years) were less likely to undergo screening compared to older women (50–64 years) [61]. This discrepancy may reflect a lower perceived cancer risk among younger adults or a tendency to prioritize acute complications of diabetes over preventive care [32]. The educational level emerged as a key predictor: 60% of women with university-level education had a recent Pap test, compared to less than 20% of those with incomplete primary education [62]. This finding aligns with global literature highlighting education as a facilitator of health literacy, health-seeking behavior, and effective system navigation [79, 43].

Furthermore, prior medical counseling was associated with a 3.5-fold increase in screening adherence, underscoring the essential role of healthcare providers in promoting preventive behavior and empowering patients during routine consultations [32].

### Comparison with the Literature

Our findings are consistent with evidence from other diabetic populations [63]. Studies from India and the United States report a 16–18% lower Pap test adherence among diabetic women compared to non-diabetics, even after adjusting for socioeconomic status [30, 2]. Similarly, the link between lower educational attainment and reduced screening is well-documented in Brazil and Colombia, where women with minimal formal education are 2.3 times more likely to forgo Pap testing [43].

Contrasts arise when compared with European cohorts, where screening rates appear uniform across age groups—likely attributable to universal health coverage, automated reminder systems, and broader availability of self-sampling kits [60]. In our cohort, obesity (present in 74% of participants) emerged as both a physical and psychological barrier [64]. While our data highlight the inadequacy of available equipment (e.g., specula, exam tables), studies from Canada and Australia emphasize the role of body image issues and provider bias in discouraging screening participation [30, 32, 31].

### Clinical and Public Health Implications

The results of this study highlight the need to rethink diabetes care models to incorporate integrated, biopsychosocial approaches to prevention [65]. The following strategies are supported by the literature and may help improve cervical cancer screening coverage [66]:

#### Systematic Integration of Pap Testing into Chronic Care

Routine DM2 follow-up appointments should systematically include reproductive health indicators, particularly cervical screening status [67]. This approach, successfully implemented in Chile’s “preventive bundle” model, has demonstrated a 30% increase in screening coverage by reducing fragmentation of care [68].

#### Tailored Health Education Interventions

Culturally and linguistically appropriate educational materials must be developed, particularly for women with limited schooling [69]. Multimedia tools, community theater, and peer-led health sessions can significantly improve knowledge and reduce stigma surrounding the Pap test [33].

#### Motivational Counseling Training for Healthcare Providers

Healthcare professionals should be trained in motivational interviewing and person-centered counseling [70]. These techniques have proven effective in addressing myths (e.g., “Pap testing is unnecessary without recent sexual activity”) and improving health engagement among underserved populations [33].

#### Infrastructure Adaptation for Obese Women

Healthcare facilities must be equipped with bariatric exam tables, larger specula, and private examination spaces [71]. Physical barriers should not prevent access to cancer screening for women living with obesity [72].

### Study Limitations

This study has several limitations inherent to its cross-sectional design [4]:

- Small sample size (*n* = 73), which limits the statistical power to detect associations in key subgroups, such as women with prior HPV infection or hysterectomy history [12].
- Recall bias, as screening history was self-reported and not validated through medical records [13].
- Institutional context, given that all participants received care through the Mexican Social Security Institute (IMSS) in an urban setting, limiting generalizability to rural or uninsured populations [7].
- Unmeasured variables, including HPV vaccination status, cervical cancer knowledge, cultural barriers (e.g., embarrassment), and access to transportation—factors previously identified as influential in screening adherence [39].

### Recommendations for Future Research

- Longitudinal studies exploring causal relationships between glycemic control (e.g., HbA1c), chronic inflammation, and cervical lesion progression [8].
- Qualitative research using in-depth interviews to explore cultural and psychological barriers to screening among indigenous women, migrants, and those living in rural areas [9].
- Randomized controlled trials comparing the effectiveness of digital health interventions (e.g., SMS reminders, mobile health apps) versus traditional community-based strategies [10].
- Cost-effectiveness analyses examining the feasibility of implementing co-testing (HPV + Pap) in diabetic populations, as recommended by current international guidelines [11].

## 5 Conclusion

This study highlights a critical gap in cervical cancer prevention among women with type 2 diabetes mellitus in a secondary-level healthcare setting in Mexico [73]. Despite their elevated risk profile, nearly half of the participants had not undergone a Pap test within the recommended interval, with disparities driven by age, educational attainment, prior counseling, and obesity-related barriers [74]. These findings underscore the need for more inclusive and integrated care models that prioritize preventive services as an essential component of diabetes management [75].

To improve screening coverage, healthcare systems should adopt patient-centered strategies, including routine screening during chronic disease follow-up, targeted health education, provider training in motivational counseling, and infrastructural adjustments that reduce physical and psychological barriers [76]. Addressing these gaps requires coordinated efforts across policy, clinical practice, and community engagement to ensure that vulnerable populations—especially women with chronic conditions—receive equitable and timely cancer prevention services [77].

Future research should further explore the sociocultural and systemic factors limiting screening uptake, while evaluating innovative interventions that bridge the divide between chronic disease care and preventive health [78].

## Acknowledgement

The authors wish to express their deep gratitude to the medical and nursing staff at Hospital General de Zona con Medicina Familiar No. 21 (HGZ/MF) for their invaluable support and collaboration throughout the development of this study. We also acknowledge the significant contribution of the academic and teaching team who thoroughly reviewed the manuscript, providing critical feedback and corrections that enhanced the scientific quality and clarity of the article. We appreciate the institutional support from the Instituto Mexicano del Seguro Social (IMSS), which facilitated the necessary resources and access for conducting this research. Finally, we extend our thanks to all individuals who, directly or indirectly, contributed to the successful completion of this work.

## Authors’ Contributions

All authors contributed equally to the conception and design of the study, data collection, data analysis, and interpretation of results. All authors participated in drafting and revising the manuscript and approved the final version for publication.

## Data Availability

The data supporting the findings of this study are available from the corresponding authors upon reasonable request. Due to privacy and ethical restrictions involving sensitive patient information, the data are not publicly available. However, anonymized datasets can be provided to qualified researchers who submit a formal request and agree to comply with relevant data protection regulations.

## Ethical Statement

This study was conducted in accordance with the ethical principles of the Declaration of Helsinki and adhered to national and institutional standards for research involving human participants [54]. Ethical approval was obtained from the local Research Ethics Committee of the ***General and Family Medicine Hospital No. 21* of the *Mexican Social Security Institute (IMSS)***. All participants were informed about the nature and objectives of the study and provided written informed consent prior to data collection. Confidentiality and anonymity of the participants were strictly maintained throughout the research process.

## Declaration Of AI Usage

No AI tools were used in the conception, data analysis, writing, or editing of this manuscript. All content was developed exclusively by the authors.

